# First assessment of a cost questionnaire for its agreement with administrative databases in females with stress urinary incontinence in Switzerland: a validation study

**DOI:** 10.1101/2023.10.30.23296075

**Authors:** Céline Moetteli, Jan Taeymans, Irene Koenig

**Affiliations:** Bern University of Applied Sciences, Department of Health Professions, Division of Physiotherapy, Bern, Switzerland; Zurich University of Applied Sciences, School of Health Professions, Institute of Physiotherapy, Winterthur, Switzerland; Vrije Universiteit Brussel, Faculty of Physical Education and Physiotherapy, Brussels, Belgium

**Keywords:** Urinary Incontinence, Stress, Data Collection, Surveys and Questionnaires, Health Care Costs

## Abstract

**Introduction:** Economic research in the field of pelvic floor rehabilitation is lacking in transparency of methodology and consideration of indirect costs. Service use questionnaires collect cost data with the broad approach needed for the societal perspective of health economic studies. Nevertheless, these questionnaires are seldom validated. Therefore, this study aimed to evaluate the accuracy of a questionnaire to assess healthcare costs of females with stress urinary incontinence (SUI) in Switzerland.

**Methods:** A questionnaire was developed to measure direct medical, direct non-medical and indirect costs in females with stress urinary incontinence living in the larger Bern area in Switzerland. Participants were recruited out of a prior study sample, where they had already completed the cost questionnaire. Administrative databases’ cost data were obtained for the investigation period. The agreement between the reported direct medical costs of the questionnaire with those from the providers’ billings was assessed using Pearson correlation coefficient (*r*), intraclass correlation coefficient (ICC) for agreement, Lin’s concordance correlation coefficient (LINCCC) and limits of agreement (LOA).

**Results:** Fourteen participants were eligible for this study. Agreement between the two methods presented a Pearson’s *r* of 0.64 [95%CI 0.16 to 1.00], ICC of 0.79 [95% CI 0.33 to 0.93] and LINCCC 0.75 [95% CI 0.19 to 0.86]. LOA results showed no evidence for systematically under- or overreporting.

**Conclusions:** The developed questionnaire seemed to be an acceptable representative for direct medical costs in females with stress urinary incontinence in Switzerland. However, direct non-medical and indirect costs were not compared to an external criterion.

## Introduction

In Switzerland, per capita healthcare expenditures rise steadily from year to year. In 2020, healthcare costs amounted 11.8% of the gross domestic product (1). In times of increasing health expenditures, it is indispensable to perform economic evaluations to ensure cost-effective resource allocation in healthcare organizations.

The estimation of costs is an essential aspect when conducting health economic analyses. Thus, valid tools are required to obtain useful cost data. Self-reported questionnaires are one alternative to gain resource utilization data (2). The societal perspective is recommended for health economic analyses, since costs shifting between sectors might be detected (3).

Hence, not only direct medical costs are taken into account, but all arising expenses concerning the macroeconomic situation (4). In contrast to health insurance administrative data, self-reported questionnaires allow this broad capture of health related costs (5). Apart from direct medical costs, direct non-medical costs such as travel expenses, costs resulting from paid or unpaid informal care or indirect costs such as absenteeism, presenteeism or early retirement are considered (6). Economic estimations based on self-reported questionnaires are, unlike medical records, not limited by lacking cooperation of administrative staff or restrictions due to privacy and security concerns. Although administrative databases only allow insight into direct medical costs, they are a rich source of accurate data. While retrospective patient reported questionnaires may be affected by recall bias, administrative data does not suffer from such influences (5). However, administrative data may be inaccessible in a time- and cost-efficient manner and inappropriate due to their limited perspective (2).

Therefore, within the scope of a cost of illness (COI) study, a questionnaire to measure health related costs in females with stress urinary incontinence (SUI) was developed. Urinary incontinence is a common problem among females, causing almost ten billion Euros of direct and indirect costs in Europe per year (7). Stress urinary incontinence is the most common type of this condition with a mean prevalence of 35% in Europe (8). In the field of pelvic floor medicine, transparency in the costing methodology and consideration of a broad perspective of costs are lacking globally as well as in Switzerland (7). Thus, the aforementioned questionnaire should be eligible for further health economic evaluations in urinary incontinence. Therefore, it ought to be published on “Database of Instruments for Resource Use Measurement (DIRUM)”; an open-access database of resource-use questionnaires for use by health economists (9).

## Methods

The aim of this study was to evaluate the questionnaire’s accuracy of assessing health related costs in females with stress urinary incontinence living in the larger area around Bern in Switzerland. Since there is no standardized method for validating health economic questionnaires, health insurance administrative data was used as the comparative value as seen in previous papers (5,6,10,11).

### Questionnaire development

The questionnaire was developed for a previous COI analysis in patients with stress urinary incontinence. However, validity of the data gathered by this tool was never evaluated. The “Health status, comorbidities and cost-of-illness in females with stress urinary incontinence living in the Canton of Bern” (12) was embedded in a randomized controlled trial investigating the effects of two pelvic floor muscle training protocols (13). The first version of the instrument was developed based on a questionnaire that has already been used in a study from the Universities of Ghent and Antwerp (Belgium), which evaluated health status and costs of obesity in adult females in Flanders (14). Face validity of the adapted cost questionnaire was operationalized by comprehensibility and completeness of its items and assessed by six experts in the field of pelvic floor rehabilitation. Additionally, the questionnaire was confirmed for comprehensibility by two pilot-test subjects. To review criterion validity in a first point, these two test subjects filled out the cost questionnaire retrospectively for the last four months and provided the health insurance bills for the same period of observation. The comparison with the survey data did not raise any major concerns, or the need for further modifications (12).

The present cost questionnaire under investigation assessed participants’ characteristics, health status and health related costs retrospectively over four months. This included all direct medical costs such as medical and therapeutic treatment costs, raises for dental services, complementary medicine, and medications. Consumed service units and price units were asked separately. In addition, direct non-medical costs (e.g., incontinence aids) and indirect costs (e.g., productivity loss such as work absenteeism and presenteeism) were assessed. “Filter questions” were used as guiding assistance for costs reporting (6). The questionnaire in full version can be found under “additional files”.

### Setting

The previous COI study of Koenig et al. (12) used a prevalence based, bottom-up approach. According to a prevalence based approach guidelines, questions of the survey targeted not only expenditures due to SUI but the entirety of healthcare expenditure (15). Out of 96 participants in the original randomized controlled trial (RCT), 37 adult female participants volunteered in the COI study and completed the cost questionnaire in an individual four-month time horizon between 2016 and 2017. Each participant returned her questionnaire during a post-intervention measurement for the RCT. The answers were then checked by the responsible examiner for completeness and missing data were completed immediately in the presence of the participants. Based on unit prices, Koenig et al. (12) estimated the total healthcare costs of the investigated four months.

The Swiss healthcare system is highly decentralized. Compulsory health insurance is offered by competing insurers with minimal annual deductibles of 300 Swiss Francs (CHF) for insured adults. Insured persons may increase deductibles up to 2’500 CHF. Insured persons also pay 10% coinsurance of these services with an annual limit of 700 CHF for adults (16). Residents are free to take out a supplementary insurance. In contrast to the compulsory health insurance, coverage of services may differ between insurance providers.

Supplementary insurances might include, for example, complementary medicine, dental treatment, or health prevention services. There are different methods of charging the fee: service providers can either invoice the insurances directly or they can bill patients, which settle up the account and claim reimbursement from the insurer (16).

### Study participants and procedure

Each participant received written information about the study design and procedures and signed the informed consent form between October 2019 and February 2020. With volunteer’s power of attorney, one of the authors (CM) was allowed insight into personal health insurances billing data of the investigated four-month period between 2016 and 2017.

The inclusion criteria for the previous randomized controlled trial were: age range 18 to 60 years, body mass index (BMI) between 18 to 30 kg/m^2^, nulliparous or at least 12 months postpartum with a negative pregnancy test, ability to read and understand the German language and to run on a treadmill. Exclusion criteria were: acute urinary tract or vaginal infections, urogenital surgery, predominant overactive bladder and urogenital prolapse over grade one pelvic organ prolapse quantification system (POP-Q). These criteria met the International Consultation on Incontinence’s recommendation for diagnosis of SUI (17).

### Statistical analysis

Characteristics of the sample were compared to the original cohort of the RCT with a Student’s *t*-test for independent samples. As a reference for the degree of incontinence, the Oxford Grading Scale was used. This scale is a classification system of the pelvic floor muscle strength where zero represents no palpable contraction and the maximum score of five signifies strong contraction (18). Median and interquartile range (IQR) of observed muscle strength values were calculated for the original and the current samples.

For comparability with other studies (19), descriptive parametric statistics were used to analyze the cost data, despite the probability of a skewed distribution. The items “medical care” and “physiotherapy” fell into the scope of compulsory health insurance. The items “dental care”, “complementary treatment” and “medication” were categorized as services within supplementary insurance. To measure criterion validity, the differences between reported costs and health insurance billings were calculated and compared with “0” difference using a one-sample *t*-test. For a first visual impression of the relationship between the datasets, a scatterplot was performed. Cost data is usually skewed due to, for example, medical complications. Therefore, data was log transformed for approximate normality to conduct calculations of correlation (20,21).

As a preliminary test, consistency of variables was indicated by Pearson’s correlation coefficient (*r*). Agreement was measured using intraclass correlation coefficient (ICC). Since the variables were continuous and reported in the same units, ICC was appropriate (22). ICC two-way, mixed model formula for agreement was applied. As a reference value for small sample sizes, Lin’s concordance correlation coefficient (LINCCC) was added (23). Pearson’s *r*, ICC and LINCCC are measurements of correlation ranging from 1 (perfect agreement) to -1 (perfect inverse agreement) (23,24). The following levels for interpreting accuracy were applied: “poor” = < 0.40, “fair to good” = 0.40 to 0.74, “excellent” = 0.75 to 1.00 (25). Standard error of measurement (SEM) was calculated (26). Systematic bias was assessed using limits of agreement (LOA) and were presented in a Bland Altman plot (6). The visual result was supported by the calculation of linear regression of the difference between the costs over mean value of the costs. The procedure was doublechecked by two of the authors independently.

All data analyses were performed using SPSS (IBM Corp. Released 2017. IBM SPSS Statistics for Windows, Version 25.0. Armonk, NY: IBM Corp.) and with a macro written in Excel for the calculation of LINCCC. Significance was set at 5% level of error.

## Results

Out of the 37 participants of the COI study, 14 persons volunteered in the present agreement study. The participants were insured in eight different healthcare insurance companies, which all provided detailed billing data after written request from the participants.

Table 1 shows the characteristics of the participants such as age, BMI, number of vaginal births as well as sectio and scores on the Oxford Grading Scale. The present sample differed in none of these variables significantly from the original RCT population (*p* > 0.05).

**Table 1.** Comparison of previous and current sample’s participants characteristics.

|  | Present study (n=14) |  | RCT (n=96) |  | *p* |
| --- | --- | --- | --- | --- | --- |
|  | Mean | SD | Mean | SD |  |
| Age (years) | 54.0 | 9.4 | 51.0 | 9.9 | 0.290 |
| BMI (kg/m<sup>2</sup>) | 24.8 | 4.6 | 24.8 | 3.8 | 0.993 |
| No vaginal birth | 1.20 | 0.90 | 1.60 | 1.10 | 0.208 |
| No sectio birth | 0.15 | 0.38 | 0.30 | 0.70 | 0.458 |
| Oxford Grading Scale (0-5) | Median: 3 | IQR: 1 | Median: 3 | IQR: 1 | - |

*SD*: Standard deviation; *IQR*: Interquartile range

In table 2, data was compiled by subgroups. Mean values and standard deviations (SD) for reported costs and billings were opposed for the items “medical care” and “physiotherapy” indicating compulsory services, and “dental care”, “complementary treatment” and “medication” representing supplementary services.

**Table 2.** Subgroup means and SDs of reported costs and billings.

|  | Reported cost (CHF) |  | Billings (CHF) |  |
| --- | --- | --- | --- | --- |
|  | Mean | SD | Mean | SD |
| Compulsory services |  |  |  |  |
| Medical care | 513.60 | 594.90 | 523.80 | 854.90 |
| Physiotherapy | 508.50 | 130.70 | 481.35 | 116.45 |
| Supplementary services |  |  |  |  |
| Dentist | 70.50 | 116.90 | 23.95 | 64.85 |
| Complementary treatment | 126.10 | 248.20 | 195.00 | 397.40 |
| Medication | 69.85 | 108.15 | 59.45 | 143.75 |

*CHF*: Swiss Francs; *SD*: Standard deviation

Table 3 depicts the analysis of correlation. Calculations were conducted for overall costs as well as expenditures of compulsory and supplementary services separately. Total sum of reported costs during the four-months period was CHF 18’038.85 and CHF 17’113. 85 for billings. Overall correlation between the reported costs and the billing sums yielded in a Pearson’s *r* of 0.64 [95% CI 0.16 to 0], ICC of 0.79 [95% CI 0.33 to 0.93] and LINCCC of 0.75 [95% CI 0.19 to 0.86].

**Table 3.**
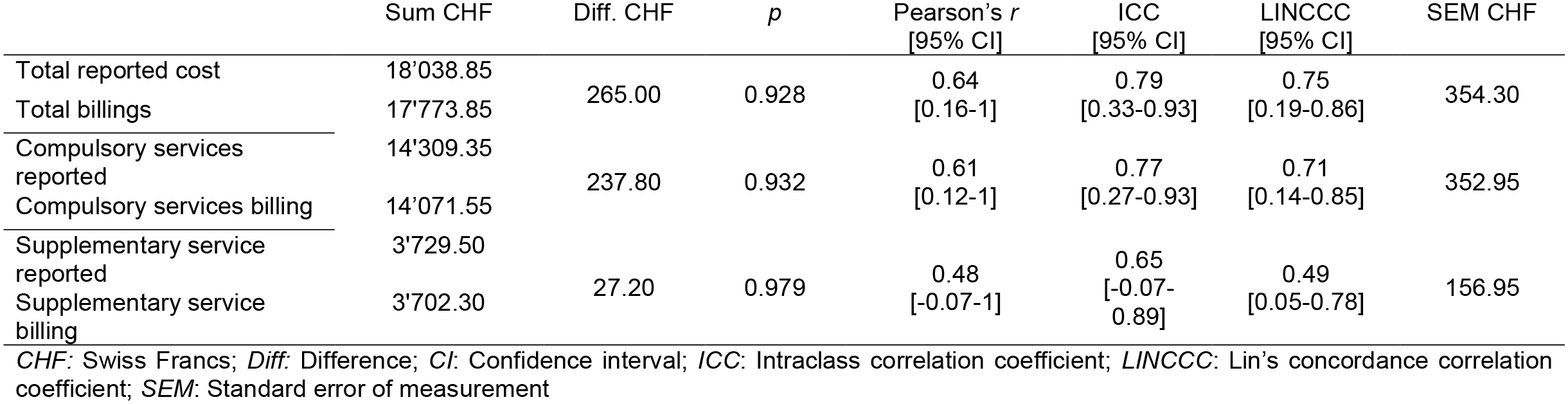
Sum, difference and correlations between reported costs and billings.

|  | Sum CHF | Diff. CHF | *p* | Pearson's *r*<br>[95% CI] | ICC<br>[95% CI] | LINCCC<br>[95% CI] | SEM CHF |
| --- | --- | --- | --- | --- | --- | --- | --- |
| Total reported cost | 18'038.85 | 265.00 | 0.928 | 0.64<br>[0.16-1] | 0.79<br>[0.33-0.93] | 0.75<br>[0.19-0.86] | 354.30 |
| Total billings | 17'773.85 |  |  |  |  |  |  |
| Compulsory services reported | 14'309.35 | 237.80 | 0.932 | 0.61<br>[0.12-1] | 0.77<br>[0.27-0.93] | 0.71<br>[0.14-0.85] | 352.95 |
| Compulsory services billing | 14'071.55 |  |  |  |  |  |  |
| Supplementary service reported | 3'729.50 | 27.20 | 0.979 | 0.48<br>[-0.07-1] | 0.65<br>[-0.07-0.89] | 0.49<br>[0.05-0.78] | 156.95 |
| Supplementary service billing | 3'702.30 |  |  |  |  |  |  |

*CHF*: Swiss Francs; *Diff*: Difference; *CI*: Confidence interval; *ICC*: Intraclass correlation coefficient; *LINCCC*: Lin's concordance correlation coefficient; *SEM*: Standard error of measurement

Systematic errors of overall data are presented in the Bland Altman Plot in Figure 1. Linear regression from cost difference over mean costs yielded a beta regression coefficient of -0.17 (*p* = 0.564).

**Figure 1.**
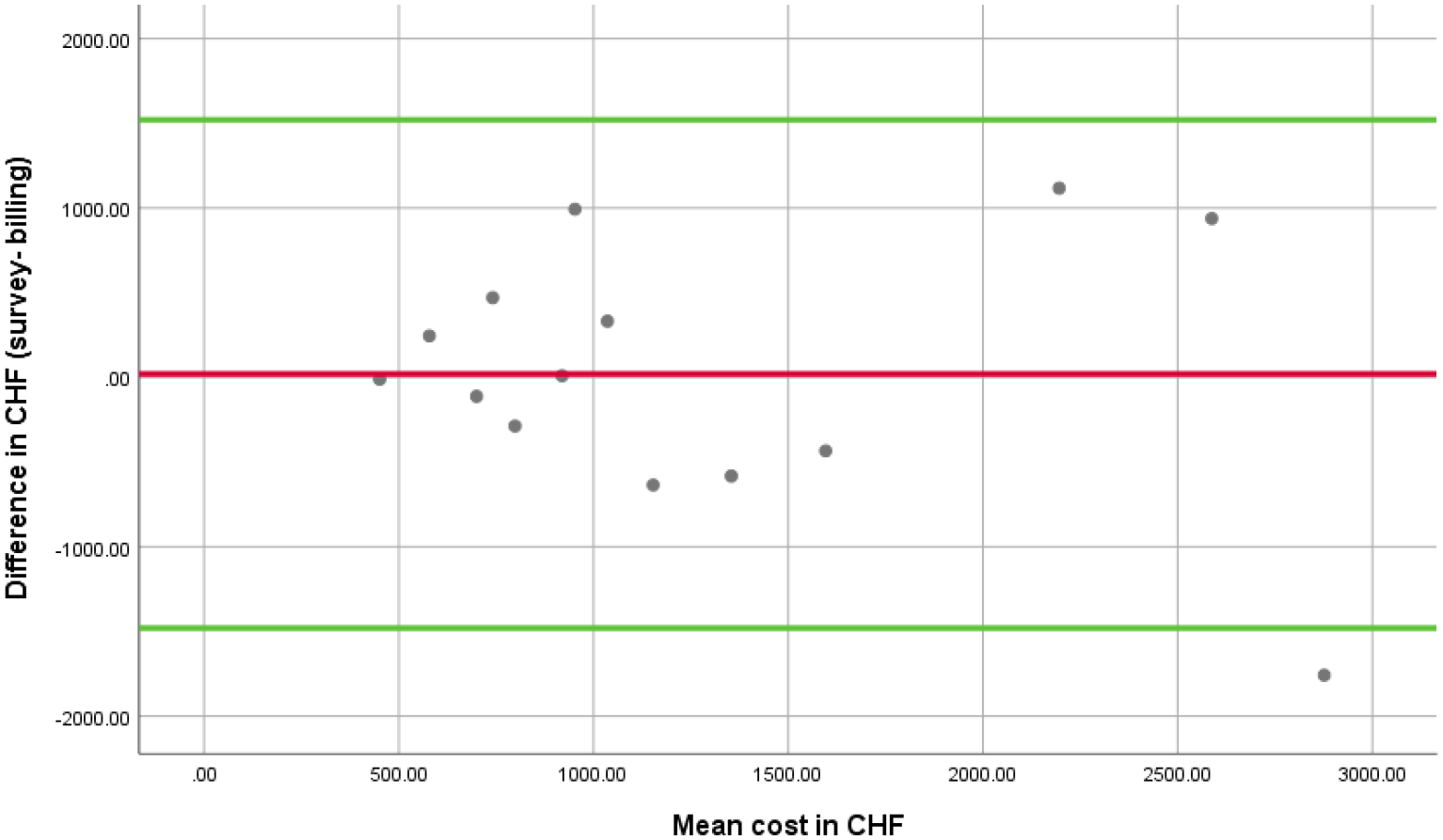
Bland Altman Plot. Mean values of provider and survey data displayed in the x-axis are plotted against the difference between the values in the y-axis. Limits of agreement are represented in green. The red line is the mean of difference and is drawn at 20.3

## Discussion

### Assessment of validity

This study aimed to validate a questionnaire assessing costs in females with stress urinary incontinence living in the larger area around Bern in Switzerland. Although analysis of correlation is known as a reliability process, it is also used as a method to determine criterion validity between the measurement instrument and the gold standard (22). As a gold standard, health insurance billing was used in this study.

Students’ *t*-test for independent samples showed no significant difference in demographic data of the 14 participants enrolled in the current study to the original cohort of 96 females. Since the participants of the RCT were diagnosed according to the criteria of the International Consultation on Incontinence, the current sample was representative for females with stress urinary incontinence.

Healthcare expenditures analyzed by the items “medical care”, “physiotherapy”, “dental care”, “complementary treatment” and “medications” showed large standard deviations in both reported and administrative data. Since each of those items represented different medical services, this variability seemed to have occurred naturally. For example, a respondent could accrue a ticket for a simple laboratory service of about CHF 20 in the category of medical care as well as a surgical procedure of CHF 1’500. Despite the observed variability, when comparing mean values of reported costs and billing data, subgroups appeared similar. This was particularly relevant for the items “medical care” and “physiotherapy”. Participants seemed to estimate costs for these services accurately and consistently with insurance billings. The differences of datasets get larger, when it comes to the costs of dental care and complementary treatments. Reported dentist costs were three times higher than insurances recorded billings. Since Swiss residents are free to take out a supplementary insurance of their needs, different services might be covered or not. If participants had a supplementary dental health insurance, dentists’ costs would be covered by the insurance. If not, residents would most likely settle the bill out of pocket without submitting the invoice to the insurance. Consequently, companies might be incompletely informed and reflect the true costs insufficiently. Thus, the patient reported questionnaire could be more accurate than the so-called gold standard itself. Concerning complementary treatments, billings were higher than reported costs. Since overestimation of billing data is unlikely (5), underestimation of patient reported cost must be assumed. Pinto et al. (5) observed this behavior of underreporting regarding doctor visits. Recall bias side lined, difference of reported costs may also be a result of the “reverse telescoping” effect.

Telescoping occurs, when participants unconsciously lengthen their period of investigation and report events erroneously into the recall time. In contrast, treatments within the time frame may be “reverse telescoped” out of the recall period (27). True costs might even be higher than insurance’ billings revealed. Depending on the coverage of services, participants may or may not have submitted the invoices to the companies. Therefore, unsubmitted billings must be assumed, which would lead to a simultaneous underestimation of insurances expenditures for complementary treatments. In the case of medications, insurance companies’ information is generally restricted to prescribed drugs. Whereas the patient reported cost questionnaire captured all types of medication, including out of pocket utilizations. Overall sums of reported costs and billings were similar but with large standard deviations. Subsequently, the data range had a similar impact on each arm of the distribution and averaged itself out. Correspondingly similar values of reported costs were found after dividing the costs into categories of compulsory and supplementary services. Differences between the data sets were nonsignificant in all the three calculations. Therefore, Pearson’s *r* correlation coefficient was calculated. In contrast to ICC agreement, Pearson’s *r* does not take systematic error into account (22). Whereas ICC measures the degree to which one variable can be equated to another variable, Pearson’s *r* assesses the linearity of the relationship between the two variables. Lin’s concordance correlation coefficient is known for its robustness even with small sample sizes (23,24). It evaluates the degree to which pairs fall on the 45° line when plotted (28). In this present study, variability between the degree of correlation between the three methods occurred. Given the small number of samples, Pearson’s *r* and ICC may have yielded unstable calculations. Hence, ICC reflected a higher correlation than Pearson’s *r* despite the more restrictive formula (22). Point estimations of correlation for overall data were “fair/ good” to “excellent” according to the levels of accuracy with a Pearson’s *r* of 0.54 [95% CI 0.16 to 1.00], ICC of 0.79 [95% CI 0.33 to 0.93] and LINCCC of 0.75 [95% CI 0.19 to 0.86]. Nevertheless, the large confidence intervals indicated an ambiguous result. As expected, due to the descriptive values, correlations of supplementary services were lower than compulsory services with point estimations rated “fair to good” for supplementary services respectively, “fair to good” and “excellent” for compulsory services.

The Bland Altman plot (figure 1) depicts the mean value of the pairs against differences between the pairs. Each dot on the plot represents data from one participant. There are six dots each above and below the mean visible. The limits of agreement are wide and thus consistent with the large confidence intervals of the correlations. Out of 14 dots, 13 are located within the limits of agreement and one is situated exteriorly. There is a trend noticeable, where the more the mean increases, the larger the differences become. This finding suggests that participants who have less health service consumption were able to estimate their costs better than those with much healthcare consumption. The calculation of the linear regression confirmed the first impression of no evident systematic bias. Beta coefficient was -0.17 (*p =* 0.564), indicating that there was no proportional bias involved.

Indeed, the plot shows that the deviations of the mean were evenly balanced between under- and overreporting of study participants. In the systematic review of Leggett et al. (2), a tendency of underreporting was noticed. The 15 included self-reported questionnaires seemed to capture the number of hospitalizations and emergency room visits accurately, whereas participants tended to underestimate the number of visits to a general practitioner (GP).

Pinto et al. (5) developed a questionnaire to capture healthcare use and costs in patients with osteoarthritis in New Zealand. This study included 50 participants. The reported costs of the three-month investigation period were compared to administrative data. GP services, hospital services and medication use were collated. According to the authors, correlations were acceptable concerning GP services and hospitalizations (LINCCC >0.4) but exhibited poor agreement for medication (LINCCC < 0.4). These results were in line with findings of the current included females with SUI. Depending on the cost categories, concordance between survey and administrative data corresponded well and the questionnaire seemed to give an appropriate reflection of direct medical costs. Nonetheless, administrative databases were limited as gold standard, since they captured a more restricted perspective of costs than the patient reported questionnaires.

This study attempted an important step for standardized economic research in the field of pelvic floor medicine and rehabilitation. Further research is needed to evaluate and strengthen the results of this present study on a greater sample size. Since the cost questionnaire is only available on paper, a digital version is required. For this future version, a feasibility study will be needed.

### Limitations

In their systematic review, Leggett et al. (2) showed that few studies have aimed to measure the accuracy of healthcare service use and cost data questionnaires. Thus, no unambiguous method is known (29) and limitations need to be declared. The first obstacle for assessing accuracy of such questionnaire is the lack of an appropriate “gold standard” (29). Insurance companies do not cover all types of healthcare costs and are therefore not informed about the totality of expenses. Due to the decentralized Swiss healthcare system, there is no single omniscient administrative database. Survey data were thus compared to a deficient criterion. Secondly, since there is no appropriate comparator for a broad perspective of costs, the cost questionnaire was assessed for a subset of questions only. Hence, declaration of validity was only possible for the part of direct medical costs. Direct non-medical and indirect costs were neglected. Thirdly, sample size of the present study was low (n = 14). Due to the fact, that the study population was originally participating in an RCT and was committed to the following COI analysis, only few people agreed with another observation. Consequently, the low number of participants may have generated unstable calculations. An unambiguous result would have been achieved by a greater cohort.

### Conclusion

The developed questionnaire for measuring costs in females with stress urinary incontinence in Switzerland seemed to capture direct medical costs sufficiently. Overall correlation of patient reported data and health insurance billings yielded in a Pearson’s *r* of 0.64 [95% CI 0.16 to 1], ICC of 0.79 [95% CI 0.33 to 0.93] and LINCCC of 0.75 [95% CI 0.19 to 0.86].

Compulsory services such as medical care and physiotherapy corresponded better with administrative database than supplementary services such as expenses for dentist, complementary treatment, and medication. No proportional bias of under- or overreporting occurred.

## Supporting information

Supplemental Questionnaire

## Data Availability

All data produced in the present study are available upon reasonable request to the authors

## List of abbreviations

BMI: body mass index
CHF: Swiss Franks
CI: confidence interval
COI: cost of illness
DIRUM: Database of Instruments for Resource Use Measurement
GP: general practitioner
ICC: intraclass correlation coefficient
IQR: interquartile range
LINCCC: Lin’s concordance correlation coefficient
LOA: limits of agreement
POP-Q: pelvic organ prolapse quantification system
RCT: randomized controlled trial
SD: standard deviations
SEM: standard error of measurement
SUI: stress urinary incontinence

## Declarations

### Ethics approval and consent to participate

The study protocol has been approved by the Ethics Committee of the Canton of Bern, Switzerland (reference number 249/14 on 27 June 2019) in accordance with the Declaration of Helsinki and the Swiss Human Research Act. Participants received written information about data collection and data protection as well as signing an informed consent form.

### Authors’ contributions

CM, IK, and JT provided input into the design of the study. CM obtained informed consent from study participants and gathered cost data from health insurers. CM and JT analyzed and interpreted the data. CM prepared the first draft of the manuscript. IK and JT reviewed the manuscript and provided input. All authors read and approved the final version of the manuscript.

## Acknowledgements

The authors would like to thank Sara Mahnig for her methodological advice, André Meichtry for his statistical support and Ben Tite for proofreading this article. Thanks also to the employees of responsible insurance companies, which compiled requested data.

## Trial registration

Baseline randomized controlled trial was registered at clinicaltrials.gov (www.clinicaltrials.gov/ study identifier: NCT02318251) in February 2018.

## Supporting information

**S1 File cost questionnaire “Health and** cost **evaluation in SUI”**.

## References

1. OECD. (24. Oktober, 2022). Anteil der Gesundheitsausgaben in der Schweiz am Bruttoinlandsprodukt von 2010 bis 2020 [Graph]. In Statista. Zugriff am 21. März 2023, von https://de.statista.com/statistik/daten/studie/18342/umfrage/anteil-der-gesundheitsausgaben-in-der-schweiz-am-bruttoinlandsprodukt-bip/l

2. Leggett LE, Khadaroo RG, Holroyd-Leduc J, Lorenzetti DL, Hanson H, Wagg A, et al. Measuring Resource Utilization: Systematic A Review of Validated Self-Reported Questionnaires. Medicine. 2016 Mar;95(10):e2759.

3. Byford S, Raftery J. Economics notes: Perspectives in economic evaluation. BMJ. 1998 May 16;316(7143):1529–30.

4. Schöffski O, Graf von der Schulenburg JM, editors. Gesundheitsökonomische Evaluationen. Berlin, Heidelberg: Springer Berlin Heidelberg; 2012.

5. Pinto D, Robertson MC, Hansen P, Abbott JH. Good agreement between questionnaire and administrative databases for health care use and costs in patients with osteoarthritis. BMC Med Res Methodol. 2011 Dec;11(1):45.

6. Schweikert B, Hahmann H, Leidl R. Development and first assessment of a questionnaire for health care utilization and costs for cardiac patients. BMC Health Serv Res. 2008 Dec;8(1):187.

7. Zwolsman S, Kastelein A, Daams J, Roovers JP, Opmeer BC. Heterogeneity of cost estimates in health economic evaluation research. A systematic review of stress urinary incontinence studies. Int Urogynecol J. 2019 Jul;30(7):1045–59.

8. Hunskaar S, Lose G, Sykes D, Voss S. The prevalence of urinary incontinence in women in four European countries. BJU Int. 2004 Feb;93(3):324–30.

9. DIRUM - Database of Instruments for Resource Use Measurement [Internet]. [cited 2020 May 8]. Available from: http://www.dirum.org/

10. Lairson DR, Basu R, Begley CE, Reynolds T. Concordance of survey and billing data in a study of outpatient healthcare cost and utilization among epilepsy patients. Epilepsy Research. 2009 Nov;87(1):59–69.

11. Goossens MEJB, Mölken MPMHR van Vlaeyen JWS, van der Linden SMJP. The cost diary. Journal of Clinical Epidemiology. 2000 Jul;53(7):688–95.

12. Koenig I, Moetteli C, Luginbuehl H, Kuhn A, Taeymans J. Health status, comorbidities and cost-of-illness in females with stress urinary incontinence living in the Canton of Bern. Zeitschrift für Evidenz, Fortbildung und Qualität im Gesundheitswesen. 2020 Apr;(150–152):73–9.

13. Luginbuehl H, Lehmann C, Baeyens JP, Kuhn A, Radlinger L. Involuntary reflexive pelvic floor muscle training in addition to standard training versus standard training alone for women with stress urinary incontinence: study protocol for a randomized controlled trial. Trials. 2015 Dec;16(1):524.

14. Hens W, Vissers D, Annemans L, Gielen J, Van Gaal L, Taeymans J, et al. Healthrelated costs in a sample of premenopausal non-diabetic overweight or obese females in Antwerp region: a cost-of-illness analysis. Arch Public Health. 2018 Dec;76(1):42.

15. Mauskopf J. Prevalence-based economic evaluation. Value In Health. 1998 Nov;Volume 1(Issue 4):Pages 251–259.

16. Mossialos E, Wenzl M, Osborn R, Sarnak D. International Profiles of Health Care Systems, 2015. The Commonwealth Fund; 2016 Jan.

17. Avery K, Donovan J, Peters TJ, Shaw C, Gotoh M, Abrams P. ICIQ: A brief and robust measure for evaluating the symptoms and impact of urinary incontinence. Neurourol Urodyn. 2004;23(4):322–30.

18. Bo K, Finckenhagen HB. Vaginal palpation of pelvic floor muscle strength: inter-test reproducibility and comparison between palpation and vaginal squeeze pressure. Acta Obstet Gynecol Scand. 2001 Oct;80(10):883–7.

19. Chernyak N, Jülich F, Kasperidus J, Stephan A, Begun A, Kaltheuner M, et al. Time cost of diabetes: Development of a questionnaire to assess time spent on diabetes self-care. Journal of Diabetes and its Complications. 2017 Jan;31(1):260–6.

20. Thompson SG. How should cost data in pragmatic randomised trials be analysed? BMJ. 2000 Apr 29;320(7243):1197–200.

21. Euser AM, Dekker FW, le Cessie S. A practical approach to Bland-Altman plots and variation coefficients for log transformed variables. Journal of Clinical Epidemiology. 2008 Oct;61(10):978–82.

22. de Vet HCW, Terwee CB, Mokkink LB, Knol DL. Measurement in Medicine: A Practical Guide. Cambridge: Cambridge University Press; 2011.

23. Lin LIK. Assay Validation Using the Concordance Correlation Coefficient. Biometrics. 1992 Jun;48(2):599.

24. McAlinden C, Khadka J, Pesudovs K. Statistical methods for conducting agreement (comparison of clinical tests) and precision (repeatability or reproducibility) studies in optometry and ophthalmology: Agreement studies. Ophthalmic and Physiological Optics. 2011 Jul;31(4):330–8.

25. Fleiss JL, Levin B, Paik MC. Statistical Methods for Rates and Proportions [Internet]. Hoboken, NJ, USA: John Wiley & Sons, Inc.; 2003 [cited 2020 May 8]. (Shewart WA, Wilks SS, editors. Wiley Series in Probability and Statistics). Available from: http://doi.wiley.com/10.1002/0471445428

26. Harvill LM. An NCME Instructional Module on. Standard Error of Measurement. Educational Measure: Issues Practice. 1991 Jun;10(2):33–41.

27. Bhandari A, Wagner T. Self-Reported Utilization of Health Care Services: Improving Measurement and Accuracy. Med Care Res Rev. 2006 Apr;63(2):217–35.

28. Lin LIK. A Concordance Correlation Coefficient to Evaluate Reproducibility. Biometrics. 1989 Mar;45(1):255.

29. Byford S, Leese M, Knapp M, Seivewright H, Cameron S, Jones V, et al. Comparison of alternative methods of collection of service use data for the economic evaluation of health care interventions. Health Econ. 2007 May;16(5):531–6.

