## Supplemental Questionnaire for "First assessment of a cost questionnaire for its agreement with administrative databases in females with stress urinary incontinence in Switzerland: a validation study"

### Health and cost evaluation

Dear participant
On the one hand, this questionnaire aims to give you an idea of your health and, on the other hand, to evaluate your health costs.
We think that urinary incontinence could be a significant cost factor for those affected, as well as for our healthcare system. Physiotherapy is the internationally accepted first line therapy. Physiotherapy is effective and cost-saving. Cost evaluation is important to show that it makes sense to prescribe physiotherapy for incontinence.

The questionnaire consists of 3 parts:
PART 1: General questions
PART 2: Questions about your health
PART 3: Questions about your health care costs

The completion of the questionnaire takes about 15 minutes.

If you have any questions about this survey, please contact Irene König:
 T: 031 848 45 24

SUIP No.: ___________ Date: ____ . ____ . ________

### Part 1: GENERAL QUESTIONS

Date of birth: ___ . ___ . ____

Number of births (vaginal = spontaneous): ____
Number of births (cesarean section): ____

Highest level of education (please tick)

- ☐ Elementary school
- ☐ Vocational school
- ☐ General qualification for university entrance
- ☐ Higher technical college
- ☐ University of Applied Sciences
- ☐ University
- ☐ PhD
- ☐ Other: ______________________________________________

Employment (please tick)

- ☐ Employed
- ☐ Unemployed
- ☐ In education
- ☐ Homemaker
- ☐ Retired
- ☐ Other: ______________________________________________

### Part 2: HEALTH RELATED QUESTIONS (EQ-5D)

Under each heading, please tick one box that best describes your health TODAY.

**Mobility**

☐ I have no problems in walking about
☐ I have slight problems in walking about
☐ I have moderate problems in walking about
☐ I have severe problems in walking about
☐ I am unable to walk about

**Self-care**

☐ I have no problems washing or dressing myself
☐ I have slight problems washing or dressing myself
☐ I have moderate problems washing or dressing myself
☐ I have severe problems washing or dressing myself
☐ I am unable to wash or dress myself

**Usual activities *(e.g. work, study, housework, family or leisure activities)***

☐ I have no problems doing my usual activities
☐ I have slight problems doing my usual activities
☐ I have moderate problems doing my usual activities
☐ I have severe problems doing my usual activities
☐ I am unable to do my usual activities

**Pain / Discomfort**

☐ I have no pain or discomfort
☐ I have slight pain or discomfort
☐ I have moderate pain or discomfort
☐ I have severe pain or discomfort
☐ I have extreme pain or discomfort

**Anxiety / Depression**

☐ I am not anxious or depressed
☐ I am slightly anxious or depressed
☐ I am moderately anxious or depressed
☐ I am severely anxious or depressed
☐ I am extremely anxious or depressed

Please tick the appropriate answer (yes / no).

Are you suffering from:

| | |
|---|---|
| Hypertonia: | ☐Yes ☐No |
| Diabetes: | ☐Yes ☐No |
| Cardiovascular diseases: | ☐Yes ☐No |
| Diseases of the gallbladder: | ☐Yes ☐No |
| Cancer: (if yes,) | ☐Yes ☐No |
| Gastrointestinal cancer: | ☐Yes ☐No |
| Uterine cancer: | ☐Yes ☐No |
| Breast cancer: | ☐Yes ☐No |
| Depression: | ☐Yes ☐No, but earlier ☐No, never |
| Insomnia: | ☐Yes ☐No |
| Arthritis: | ☐Yes ☐No |
| Osteoarthritis: | ☐Yes ☐No |
| Back or neck problems: | ☐Yes ☐No |
| Joint problems: | ☐Yes ☐No |
| Incontinence: | ☐Yes ☐No |
| Stress urinary incontinence: | ☐Yes ☐No |
| Urge incontinence: | ☐Yes ☐No |
| Fecal incontinence | ☐Yes ☐No |
| Prolapse (uterus, bladder) | ☐Yes ☐No |
| Removal of ovaries: | ☐Yes ☐No |
| Removal of uterus: | ☐Yes ☐No |

Are you suffering from:

| | |
|---|---|
| Allergies: | ☐Yes ☐No |
| Asthma: | ☐Yes ☐No |
| COPD (chronic lung disease): | ☐Yes ☐No |
| Skin diseases: | ☐Yes ☐No, but earlier ☐No, never |
| Thyroid under- / over-function: | ☐Yes ☐No |
| Inguinal hernia: | ☐Yes ☐No, but earlier ☐No, never |
| Gastric ulcer: | ☐Yes ☐No, but earlier ☐No, never |
| Cirrhosis of the liver: | ☐Yes ☐No |
| Chronic hepatitis: | ☐Yes ☐No |
| Kidney stones: | ☐Yes ☐No |
| Migraine: | ☐Yes ☐No |
| Parkinson's disease: | ☐Yes ☐No |
| Epilepsy: | ☐Yes ☐No |
| HIV / AIDS: | ☐Yes ☐No |

Other diseases: ____________________________________________________________

### PART 3: Questions about your health care costs

The aim of the following questions is to know the general medical costs (consultations, medications, examinations, etc.) you had during the last four months of the intervention phase. If you are not sure, you can estimate the costs.
Please mark with a question mark (?) if you estimated the number, time or cost of the consultations.

**How many times have you consulted your doctor over the past four months?**
Number of consultations: ____
Average time per consultation: ____ min

What did these consultations cost in total?
Total: ________ CHF

Who paid for it?
☐Private ☐Health insurance ☐Other: ________________________________

**How many times have you consulted a specialist during the past four months?**
Number of consultations: ____
Average time per consultation: ____ min

What did these consultations cost in total?
Total: ________ CHF

Who paid for it?
☐Private ☐Health insurance ☐Other: ________________________________

**How many times have you received help from SPITEX in the past four months?**
Number of consultations: ____
Average time per consultation: ____ min

What did these consultations cost in total?
Total: ________ CHF

Who paid for it?
☐Private ☐Health insurance ☐Other: ________________________________

**How many times have you taken an alternative medical consultation (homeopathy, massages, kinesiology, etc.) in the last four months?**
Number of consultations: ____
Average time per consultation: ____ min

What did these consultations cost in total?
Total: ________ CHF

Who paid for it?
☐Private ☐Health insurance ☐Other: ________________________________

**How many times have you consulted a physical therapist during the past four months?**
Number of consultations: _____
Average time per consultation: _____ min

What did these consultations cost in total?
Total: __________ CHF

Who paid for it?
☐Private ☐Health insurance ☐Other: ________________________________________

**How many times have you consulted an ergo-therapist during the past four months?**
Number of consultations: _____
Average time per consultation: _____ min

What did these consultations cost in total?
Total: __________ CHF

Who paid for it?
☐Private ☐Health insurance ☐Other: ________________________________________

**How many times have you consulted a dentist during the past four months?**
Number of consultations: _____
Average time per consultation: _____ min

What did these consultations cost in total?
Total: __________ CHF

Who paid for it?
☐Private ☐Health insurance ☐Other: ________________________________________

Please tick the appropriate answer (yes / no). If your answer is yes, then please complete the following table.

**Have you taken any medications in the last four months?**

☐Yes ☐No

If yes, please complete the following table:

| Medications (Name) | Reason | Ingest: No of days in the last for months | Amount per day | Price per packet | No of pills per packet |
|---|---|---|---|---|---|

**Have you undergone medical examinations in the last four months (X-rays, blood samples etc.)?**

☐Yes ☐No

If yes, please complete the following table:

| What kind of examination | Reason | No of examinations in the last four month | Costs (hospital bill, doctor, ...) |
|---|---|---|---|

**Have you been hospitalized in the last four months?**

☐Yes ☐No

If so, how many nights have you been in the hospital for the last four months?
Number of nights: ______

**Have you had surgery in the last four months?**

☐Yes ☐No

If yes, please complete the following table:

| What kind of surgery | Reason | Costs (hospital bill) |
|---|---|---|

**Have you been in a day clinic / emergency for the past four months?**

☐Yes ☐No

If yes, please complete the following table:

| Name of the hospital | Reason | Costs (hospital bill) |
|---|---|---|

**Have you used pads for your incontinence (in the past four months)?**

☐Yes ☐No

Number of sanitary napkins in four months: ____

Total costs in four months?
Total: ________ CHF

Who paid for it?
☐Private ☐Health insurance ☐Other: ________________________________

**Have you had any extra costs for more laundry or clothes because of your incontinence (in the past four months)?**

☐Yes ☐No

Number of sanitary napkins in four months: ____

Total costs in four months?
Total: ________ CHF

Who paid for it?
☐Private ☐Health insurance ☐Other: ________________________________

**Have you had any extra costs for transportation because of your incontinence (in the past four months)?**

☐Yes ☐No

Total costs in four months?
Total: ________ CHF

Who paid for it?
☐Private ☐Health insurance ☐Other: ________________________________

**Have you had less working time because of your incontinence (in the past four months)?**

☐Yes ☐No

If so, the loss of work has affected the following areas (multiple answers possible):

☐Self-employed
☐Employed
☐Voluntary work
☐Housekeeping
☐Other: ______________________________________________________________

Total: ________ Days

**Was your efficiency limited during work because of your incontinence (in the past four months)?**

☐Yes: _________ % loss of efficiency
☐No

**Was your physical activity limited because of your incontinence (free time, sports etc.)?**

☐Yes: _________ total hours
☐No

**Was your social contact limited because of your incontinence (free time, sports etc.)?**

☐Yes: _________ total hours
☐No

**Have you had other than the costs already mentioned because of your incontinence (in the last four months)?**

☐Yes ☐No

If so, what kind of costs?

_______________________________________________________________________________

Total costs: _________ CHF

**Were you on average psychologically stressed because of your incontinence (in the last 4 months)?** (Please tick clearly!)

How strong was the load on a scale of 0 - 10?
(0 = not loaded, 10 = maximum loaded)

☐0 ☐1 ☐2 ☐3 ☐4 ☐5 ☐6 ☐7 ☐8 ☐9 ☐10

**Have you ever been asked by a doctor if you have any incontinence problems?**

☐Yes ☐No

If so, when was it the first time?
___ / ____ (Month / Year)

**Have you been informed by a doctor about the possibility of physiotherapy for incontinence?**

☐Yes ☐No

If so, when was it the first time?
___ / ____ (Month / Year)
